# Probing Gut-Brain Links in Alzheimer’s Disease with Rifaximin

**DOI:** 10.1101/2021.11.22.21266123

**Authors:** Paul V. Suhocki, James S. Ronald, Anna Mae E. Diehl, David M. Murdoch, P. Murali Doraiswamy

**Author notes:** Corresponding Author: Paul V. Suhocki, M.D.

## Abstract

Gut-microbiome-inflammation interactions have been linked to neurodegeneration in Alzheimer’s disease (AD) and other disorders. We hypothesized that treatment with rifaximin, a minimally absorbed gut-specific antibiotic, may modify the neurodegenerative process by changing gut flora and reducing neurotoxic microbial drivers of inflammation. In a pilot, open-label trial, we treated 10 subjects with mild to moderate probable AD dementia (MMSE = 17 ± 3) with rifaximin for 3 months. Treatment was associated with a significant reduction in serum neurofilament-light levels (p <0.004) and a significant increase in fecal phylum Firmicutes microbiota. Serum pTau181 and GFAP levels were reduced (effect sizes of -0.41 and -0.48 respectively) but did not reach significance. There was also a non-significant downward trend in serum cytokine IL-6 and IL-13 levels. Increases in stool Erysipelatoclostridium were correlated significantly with reductions in serum pTau 181 and serum GFAP. Insights from this pilot trial are being used to design a larger placebo-controlled clinical trial to determine if specific microbial flora/products underlie neurodegeneration, and whether rifaximin is clinically efficacious as a therapeutic.

**Research in Context:** *Systematic Review:* PubMed reviews showed emerging evidence for gut-microbiome-inflammation links in Alzheimer’s disease (AD).

*Interpretation:* Our pilot study revealed that rifaximin, a minimally absorbed, gut-specific antibiotic, reduced surrogate markers of neurodegeneration while increasing, potentially beneficial, microbiota in phylum Firmicutes. These data provide initial support to the hypothesis that microbiome related products may play a role in neurodegenerative disorders.

*Future Directions:* We plan to conduct additional human and pre-clinical studies to confirm these findings and determine the potential of rifaximin as a therapeutic for AD.

## Narrative

Accumulating evidence links alterations in the brain-gut-microbiome axis to Alzheimer’s disease (AD) and other neurodegenerative disorders (1-5). However, the exact causal role of the microbiome in the neurodegenerative process remains unknown. Human gut microbiota are predominantly composed of bacteria from three major phyla, namely *Firmicutes, Bacteroidetes*, and *Actinobacteria* (1). These organisms may harbor 50- to 100-fold more genes, compared to the host, and can subtly alter many aspects of human physiology.

One hypothesis gaining traction is that many different types of infections may lead to AD via a common pathway (6). In this hypothesis, microbial cells/fragments or toxic products emanating from the gut microbiome may enter the aging brain and trigger inflammation and neurodegeneration. Support for this hypothesis comes from both pre-clinical and clinical studies. For example, clinical studies suggest there are blood-brain barrier changes with aging and accumulation of bacterial endotoxin lipopolysaccharide (LPS) in the hippocampus (7). Blood levels of LPS have been reported to be higher in AD (8) and associated with amyloid pathology (9). There are also associations between neurotoxic bile acids (products of gut bacterial metabolism) and AD biomarkers (10). In addition, neuroprotective bile acids have shown promise in the treatment of ALS, another neurodegenerative disorder (11).

We hypothesized that reducing exposure to neurotoxic bacterial products (such as LPS or certain bile acids) would reduce neuroinflammation and neurodegeneration in AD. One potential approach to probe such links is through the use of antibiotics or probiotics. For example, colonization of germ-free APP-PS1 mice with microbiota from aged wild type mice leads to increased cerebral AD pathology (12) and treatment with an antibiotic reduces such pathology (13). We identified rifaximin as a drug that may be well suited to test this hypothesis in AD. Rifaximin is a gut-selective antibiotic that binds to the beta-subunit of the bacterial DNA-dependent RNA polymerase (14). Animal models, culture studies and metagenomic analyses have demonstrated that rifaximin improves the integrity of the gut epithelial barrier (15) and decreases virulence, motility and adhesion of pathogens (16). Rifaximin has also been shown to down-regulate epithelium damaging metalloproteinase and to increase the abundance of beneficial gut bacteria such as Lactobacillus (17). It has a relatively good safety profile, lack of drug interactions, and oral dosing. Human studies have demonstrated neurological benefits for treating and preventing the recurrence of hepatic encephalopathy (18). Hence, we repurposed it for use in AD and other neurodegenerative disorders.

In a pilot biomarker trial, we treated 10 subjects with mild to moderate AD (MMSE 17.3 +/- 3.3) for 12 weeks with rifaximin 500 mg bid. Rifaximin treatment was associated with a significant reduction in serum neurofilament-light (NfL) levels as well as non-significant reductions in serum tau and inflammatory cytokines. Although our trial was too short to detect cognitive benefits, NfL is a biomarker for axonal neurodegeneration and hence reductions in NfL may potentially be indicative of a future cognitive benefit. Rifaximin treatment in our trial also significantly and selectively increased the abundance of microbiota in seven genera of the phylum Firmicutes – with increases in one of these genera also significantly correlating with reductions in pTau181 and GFAP. Firmicutes may play a vital role in multiple gut and brain functions such as preserving the gut integrity (through synthesis of short chain fatty acids), neurotransmitter metabolism, polyphenol absorption, and reducing risk for microbial toxicity (5, 19-23). Prior studies have noted that the abundance of firmicutes is reduced in AD (19-25) and hence our findings also suggest a potential neuroprotective effect. Rifaximin was relatively well tolerated, with transient diarrhea being the most common side effect.

Overall, our study provides preliminary support to the hypothesis that microbiome modulation can alter markers of neurodegeneration. Because of the limitations of a small pilot study, there are still many unknowns including the mechanisms involved and the clinical significance of the observed changes. We are planning a larger (N=100, 12 month), placebo-controlled study to more definitively examine the effects of rifaximin on microbiome composition and toxicity (by measuring neurotoxic products such as LPS and bile acids) in relation to neuropathological and cognitive changes.

## Consolidated Results and Study Design

This study was approved by the Duke IRB and all subjects/legal representatives gave written informed consent (Clinicaltrials.gov identifier NCT03856359). An IND exemption was obtained for the study. 10 subjects with probable mild to moderate AD were recruited. Key inclusion criteria were MMSE score of 10-23 and stable health; key exclusion criteria were recent history of antibiotic use, cirrhosis and diarrhea. All subjects were treated with rifaximin 500 mg twice daily for 3 months. Following the end of treatment, subjects were monitored for an additional month for any adverse effects.

Serum samples at baseline and at 3 months were analyzed for neural (NfL), pathological (total tau, pTau181) and glial (glial fibrillary acidic protein (GFAP) markers as well as cytokines. Baseline and post-treatment fecal samples underwent microbiome analyses. Serum biomarker analysis was done using ultrasensitive SIMOA assay (Quanterix, Billerica, MA) whereas cytokines were assayed using high sensitivity T-cell panel (MilliporeSigma, Saint Louis, MO). Baseline and post-treatment fecal samples underwent microbiome 16s sequencing using the Qiagen PowerSoilPro DNA Kit (Qiagen, Germantown, MD). Bacterial community composition was characterized by amplification of the V4 variable region of the 16S rRNA gene by polymerase chain reaction using the forward primer 515 and reverse primer 806 following the Earth Microbiome Project protocol. Concentrations of the PCR products were accessed using a Qubit dsDNA HS assay kit (ThermoFisher, Waltham MA) and a Promega GloMax plate reader. Equimolar 16S rRNA Polymerase Chain Reaction products from all samples were pooled prior to sequencing. Sequencing was performed by the Duke Sequencing and Genomic Technologies shared resource on a MiSeq (Illumina, San Diego, CA) instrument configured for 250 base-pair paired-end sequencing runs. Raw reads were processed into Amplicon Sequence Variant count tables via the Qiime2 pipeline (2020.2). Raw sequence data was demultiplexed and quality filtered using the q2-demux plugin followed by denoising with Divisive Amplicon Denoising Algorithm 2. Reads were trimmed at the beginning or truncated at the end if the median base quality fell below a score of 30 as determined by the fastx quality stats tool from Fastx-toolkit (v0.0.14). All amplicon sequence variants are aligned with Mafft (via q2-alignment) and used to construct a phylogeny with RAxML version 8 (via q2-phylogeny). Taxonomy was assigned to Amplicon Sequence Variants using the q2-feature-classifier classify-sklearn naïve Bayes taxonomy classifier against the SILVA 132 database. Changes from baseline to endpoint (intent to treat) in biomarker and cognitive test scores were analyzed using two-tailed paired t-tests and the Spearman test examined correlations (Rv3.5.2).

Ten subjects (9 F, mean [SD] age 72.5 ± 5.8 years) with mild to moderate AD dementia (MMSE 17.3 ± 3.3) were enrolled. Serum NfL levels were significantly reduced following rifaximin treatment (p<0.004, effect size=-1.21) (Figure 1). pTau 181 and GFAP levels were reduced (effect sizes of -0.41 and -0.48 respectively) but did not reach significance (p=0.23 and p=0.17 respectively). Serum cytokine levels were lowered non-significantly for IL-6 (−1.62, p=0.07) and IL-13 (−1.78, p=0.09). The mean changes for IL-1beta (−0.1, p=0.65), IL-2 (−0.33, p=0.18), IL-4 (−17.4, p=0.44), IL-5 (−0.87, p=0.20), IL-8 (−1.31, p=0.53), IL-10 (−0.70, p=0.70), TNF-alpha (−0.46, p=0.36) were not significant. The mean 12-week changes in ADAS-Cog-11 (0.7, p =.60) and MMSE (−0.3, p = 0.67) were not significant. There was a significant increase in 7 genera of the phylum Firmicutes (Figure 2). The latter included Lactobacillus, Erysipelatoclostridium, Faecalitalea, Erysipelotrichaceae, Anaerostipes, Blautia and Ruminiclostridum. There was a significant inverse correlation between increases in Erysipelatoclostridium and reduction in pTau 181 (Rho = -0.883, p = 0.003) and GFAP (Rho = –0.35, p 0.036). Rifaximin was relatively well tolerated, with the most common side effect being transient diarrhea (N=4). One subject discontinued early due to worsening dementia. There were no serious adverse events.

**Figure 1.**
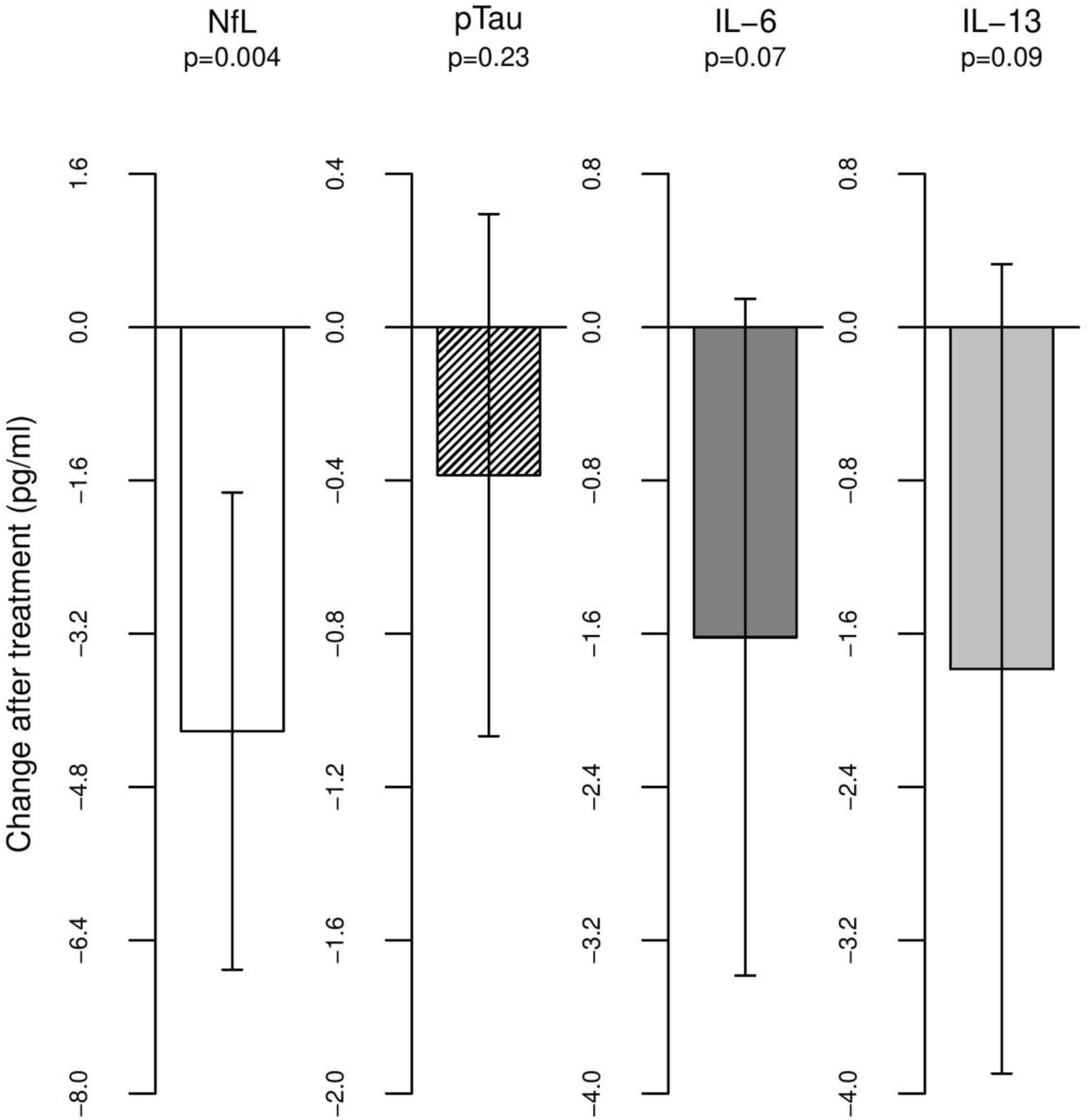
Mean (SD) reductions from baseline in serum NfL, pTau181 and cytokines.

**Figure 2.**
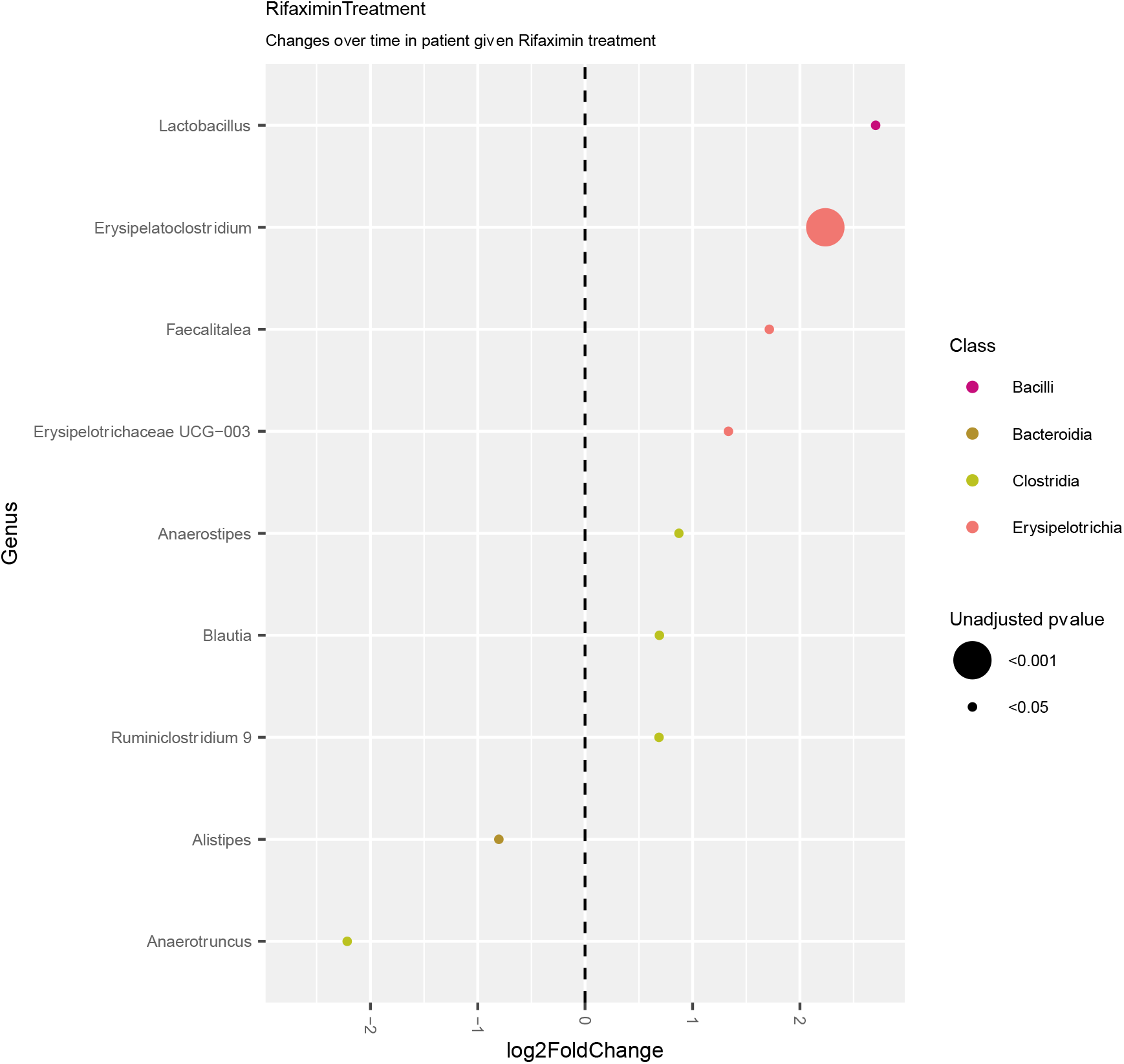
Significant increases in the abundance of 7 genera of Phylum Firmicutes following rifaximin treatment. Lactobacillus p 0.019, Erysipelatoclostridium p 0.00035, Faecalitalea p 0.02, Erysypelotrichaceae p 0.0008, Anaerostipes p 0.036, Blautia p 0.004, Ruminoclostridium 9 p 0.027

## Discussion

There are relatively few gut microbiome modulating drugs in the late stage pipeline for treatment of AD dementia. Of the 17 disease modifying therapies in Phase 3 clinical trials, only two are known to directly target the gut-brain axis (26). Atuzaginstat is a bacterial protease inhibitor that targets gingipain, a Porphyromonas gingivalis byproduct, that has been associated with neuroinflammation and hippocampal degeneration. Sodium oligomannate is an algae derived oligosaccharide that alters the gut microbiome to reduce peripheral and central inflammation. A number of other candidates are in early stage development including probiotics, bile acids, neuroprotective microbes and antibodies against toxic microbial polysaccharide antigens.

We repurposed rifaximin for use in AD because of its high gut-specificity and favorable safety profile over decades of use (18). Rifaximin is US FDA approved for the treatment of hepatic encephalopathy, travelers’ diarrhea and inflammatory bowel disease. Less than 0.4% of this antibiotic is absorbed (27), making it well-tolerated for treating gastrointestinal infections. It reduces the virulence of gut pathogens (28) and significantly lowers blood LPS levels (29). Rifaximin may also fortify the host gut epithelial barrier by blocking the ability of pathogens to attach to the gut epithelial cell surface or to be internalized by the epithelial cells (30) (31). This significantly reduces translocation of pathogens from the gut lumen into the lamina propria and mesenteric lymph nodes (32). Rifaximin, via gut epithelial cell pregnane x receptor (PXR), may also promote the transcription of genes for detoxification enzymes and cytokines (33, 34). These mechanisms have the potential to benefit not just AD but other neurodegenerative disorders.

Our preliminary findings support the hypothesis that bacterial products may contribute to neurodegeneration and that rifaximin could prove useful to probe and potentially treat these underlying mechanisms. The significant lowering of NfL levels along with decreases in serum IL-6 support the hypothesis that neuroinflammatory changes may mediate microbiome effects on neurodegeneration. Prior studies have noted increased expression of IL-6 in AD brains prior to the development of neuritic plaques suggesting inflammation may be a cause rather than a response (35) (9).

There are several microbial products that could potentially lead to neurotoxic or neuroinflammatory effects in AD. This includes ammonia (36), LPS (8), and secondary bile acids(10). Bacterial products can cross the gut epithelial barrier into the lamina propria and gain access into the brain via either the vagus nerve (37) or a compromised blood-brain barrier (38). We did not detect any changes in blood ammonia in our study suggesting that in subjects with healthy livers this may not be a major contributor. LPS, a cell wall component of gram negative gut pathogens (39), is elevated in the blood and brains of AD patients (40) (7, 41) and may induce Aß production and myelin destruction through cytokines (40). In mouse models, Kahn et al showed that peripheral LPS-induced inflammation can cause cognitive deficits and significantly increased hippocampal beta amyloid (42). We did not measure LPS but plan to do so in our next study.

Microbiota in the Firmicutes genera (Lactobacillus, Erysipelatoclostridium, Anaerostipes and Blautia) were significantly increased in abundance in our patients following treatment. Prior studies have noted a lower abundance of fecal Firmicutes in AD and this correlated with elevated CSF biomarkers of neurodegeneration (5). Firmicutes genera have previously been reported to have neuroprotective effects involving multiple mechanisms. Lactobacillus is involved in production of neurotransmitters and metabolites, including GABA, serotonin, acetylcholine, histamine, dopamine and short chain fatty acids (19). Erysipelatoclostridium promotes gut polyphenolic compound absorption (43) which can cross into the brain and protect against nitric oxide production and pro-inflammatory cytokines (44). Polyphenolic compounds are also anticholinesterase inhibitors and may have direct cognitive benefits (45). A prior study found a correlation between abundance of Erysipelatoclostridium in the gut and cognitive function in APP/PS1 mice (24). Anaerostipes production of butyrate helps to maintain a healthy gut epithelial barrier by fueling enterocytes (46). Based on these data, we have refined our hypothesis that rifaximin may also have potential probiotic effects through increasing the abundance of beneficial neuroprotective Firmicutes strains. Our findings are also supported by two additional studies (47-48). In a trial of 15 patients with irritable bowel syndrome, rifaximin increased bacterial diversity, the Firmicutes/Bacteroidetes ratio and the abundance of Faecalibacterium prausnitzii, a butyrate producer with strong anti-inflammatory properties (47). In another report on 20 patients with a variety of gastrointestinal and liver disorders, rifaximin increased the abundance of Lactobacilli (48). Additional studies in larger samples are needed to test this hypothesis and determine if the microbial changes correlate with biomarker/clinical benefits. Animal models may also help elucidate mechanisms involved.

Limitations in our study include open design, short duration and multiplicity of tests (Type 1 error) which may reduce the likelihood that our findings reflect a true effect. We also did not strictly control for diet or time of stool collection and hence our fecal microbiome analyses should be viewed as preliminary. There also remain many unknowns in the microbial hypothesis of AD, such as the exact microbial organisms and/or products implicated in neurodegeneration, how they enter the brain to cause neurodegeneration, whether biomarker changes observed (e.g. NfL) translate into meaningful clinical benefits as well as the optimal dosing of rifaximin in AD and other neurodegenerative disorders.

Despite such limitations, to our knowledge, this is the first study to demonstrate that microbiome modulation alters a marker of neurodegeneration in human AD subjects. Our intent was to generate pilot biomarker data to refine our hypothesis and power a larger placebo-controlled study to more definitively examine the effects of rifaximin on microbial composition, microbial products (e.g. LPS) and neurodegeneration markers along with cognitive and safety measures. Since rifaximin is already approved for other indications, we plan to conduct the next study also under an IND exemption. While we plan to retain the same dosing in the next study, additional dose finding studies are needed to assess whether lower doses or intermittent dosing is safer for long-term use. Because the drug is relatively free of drug-drug interactions it can also be used with existing AD therapies in combination. We are also exploring whether the specific microbes identified in our study could be developed into a therapeutic for AD. Lastly, our findings may also have relevance for other neurodegenerative disorders such as MCI, ALS and PD and studies may be warranted in those disorders.

## Data Availability

All data produced in the present study are available upon reasonable request to the authors

